# Gender Differences in Work Expectations and Psychological Distress: Insights from the United States and South Korea

**DOI:** 10.1101/2025.01.30.25321419

**Authors:** Linh Dang, Toni Antonucci, Carlos Mendes de Leon, Briana Mezuk

## Abstract

**Objectives:** Over the past 50 years, women’s roles have changed significantly, from primarily domestic roles to active labor force participation. Women from the “Baby Boom” cohort (born 1948-1965) have experienced unprecedented access to educational and employment opportunities, which has implications for work and mental health later in life. This study examined gender differences in the association between work expectations and psychological distress among Baby Boomers in the United States and South Korea.

**Methods:** Data came from the 2006-2018 waves of the Health and Retirement Study (n=14,005) and Korean Longitudinal Study of Aging (n=2,362). Perceived expectations of working in the next five years were reported on a probability scale (0-100%). Psychological distress was assessed by the Center for Epidemiologic Studies Depression Scale. Multivariate mixed-effects logistic regression models were used to examine the longitudinal association between work expectations and psychological distress for men and women.

**Results:** In both countries, women had higher CES-D scores and lower expectations of working in the next 5 years than men. Higher work expectations were robustly associated with lower odds of psychological distress among both men and women in Korea; however, this inverse association was more modest in the US. Moreover, the association between work expectations and distress was stronger among Korean men (OR_men_=0.82 [0.78, 0.87]) compared to women (OR_women_=0.88 [0.83, 0.93]), but there were no gender differences among US respondents (OR_men_=0.99 [0.96, 1.01]; OR_women_=0.96 [0.95, 0.98]).

**Discussion:** Findings highlight the complex interplay between biology sex and social contexts in shaping expectations regarding work in later life.

## INTRODUCTION

Among the most recognizable social movement in the 20th century, the women’s rights movement significantly transformed the lives of the “Baby Boom” generation (born 1948 – 1965), a cohort who entered adulthood in the late 1960s and early 1970s (Agree, 2017). The movement empowered this generation of women to defy traditional gender attitudes in an era when their primary social expectations were to be the homemakers and caregivers of their families. Women from the Baby Boom generation had unprecedented access to educational and employment opportunities; indeed, they are the first to achieve higher rates of college completion than men (Bailey & DiPrete, 2016). In addition to increasing access to education, women of this cohort have also entered the workforce in historic numbers since the 1960s. In the United States (US), the labor force participation rate of the Baby Boom women increased dramatically, from 38% in 1960 to 43% in 1970, and reached 60% by 2000 (Toossi, 2002). Together, these educational and employment gains have brought a notable shift in the roles of women in society.

### Implications of women’s labor force participation for employment in later life

Participation in the workforce has both positive and negative implications for working women as they age. On the positive side, work provides personal fulfillment and greater economic freedom as women become less dependent on their partner for financial support (Richardson, 1999). For example, retired women from the Baby Boom generation are the first who receives pension incomes based on their own earnings in addition to, rather than solely as, spousal benefits based on their partners’ earnings. In 2014, the Social Security Administration reported that more than 70% of American women received retirement benefits based on their earnings rather than marriage-based benefits (Iams, 2016). However, despite increasing access to paid work outside the home, women’s responsibilities in the domain of family life have been slower to change. Among married couples where both members work, women are still the primary caregivers of the household (Pacheco Barzallo et al., 2024), and women spend more time on housework and caregiving activities than men (Langner & Furstenberg, 2020). They are more likely to take leaves from work or take longer leaves for child and elder family care (Herr et al., 2020). As a result, women often juggle multiple roles as employees, spouses, mothers, and caregivers (Spurlock, 1995).

Among dual-earner households, competing demands between multiple work and family roles often lead to work-family conflict, and such conflict influences employment behaviors differently for men and women. In particular, men’s workforce participation typically follows a linear employment trajectory (e.g., full-time employment to complete retirement) (Richardson, 1999). In contrast, women may transition between various career paths, from full-time employment to intermittent work such as part-time employment or exiting the workforce for child or elder care responsibilities (Damaske & Frech, 2016). For example, in a study of British civil servants, older female workers with higher levels of family-to-work conflict were more likely to exit the workforce, whereas male workers who perceived greater family-to-work conflict were less likely to exit work (Xue et al., 2020). Beyond the actual work transitions, work-family conflict is related to the expectations of working in the future; prior studies show that women with higher levels of family-to-work conflict have stronger preferences for retirement than men (Raymo & Sweeney, 2006). Together, these findings demonstrate that men and women navigate work and family responsibilities differently, with stronger emphasis on family roles among women than men (Noor, 2004).

### Differences in work-related psychological distress among men and women

Extensive research demonstrates that the exposure to, and experience of, workplace stressors differ across men and women (Biswas et al., 2021). As a prime example, the wage gap is a widely known gender inequality relating to employment, with women earning significantly less than men even when both work for the same employer or in the same occupation (Penner et al., 2023). This gender wage gap increases with age; for example, American women ages 55-64 earn only 80% as much as men of the same ages, in comparison to a ratio of 92% among workers ages 25-34 (Kochhar, 2023). In addition to wage gap, gender inequalities in employment opportunities (e.g., higher prevalence of unpaid or part-time work among women) also contribute to a higher risk of psychological distress among older women compared to men (Gueltzow et al., 2023). Lastly, there are gender differences in the types of psychosocial exposures at the workplace. Specifically, men are more likely to work in physically demanding occupations and at a higher risk for workplace injuries, whereas women are more likely to experience discrimination and harassment which may elevate their risk for poor mental health (Biswas et al., 2021). Over the life course, these differences can result in differences in the relationships between work and health of older men and women (Ervin et al., 2022).

### Gender norms and employment expectations in cross-country context

Beyond geopolitical boundaries, countries vary across many contextual features that may shape work expectations and mental health. Among these include cultural values (e.g., individualism versus collectivism), gender norms and expectations (e.g., pro-women’s rights and gender equality), and workplace policies (e.g., anti-discrimination laws). For example, cultural values influence how individuals experience and navigate work-family conflict within a given culture. Work demands are strongly correlated with work-interference-with-family conflicts in individualism culture such as the US (Spector et al., 2007). In contrast, family demands are correlated with family-interference-with-work conflicts in collectivist culture (Yang et al., 2000); thus, individuals in these societies are more interested in part-time employment to fulfill their family responsibilities (Oishi et al., 2015). Moreover, gender norms also shape the workforce participation for men and women. For instance, in Asian countries, more entrenched gender inequality norms may result in discriminatory employment practices which limit women’s opportunities in the workforce (Cislaghi et al., 2022). Finally, workplace and family-related policies, including equal pay, anti-discrimination laws, and childcare benefits, can influence employment decisions, especially for female workers (Aycan, 2008; Bailey & DiPrete, 2016).

These findings reflect the social and cultural differences in work and family roles across countries, suggesting opportunities for cross-country comparisons to gain insights into the contextual factors that may shape work and mental health later in life.

### Present study

While previous research showed that the subjective expectations regarding future employment were associated with mental health among older adults (Abrams et al., 2021; Mezuk et al., 2022), it is unclear whether this association differs for men and women. The purpose of this study was to examine gender differences in the association between work expectations and psychological distress over a 12-year period. The study focused on a sample of older adults from the “Baby Boom” cohort (born 1948 – 1965) because of their unique labor force history (i.e., increasing proportion of women in the workforce). Moreover, the association between expectations and psychological distress were explored in the US and South Korea (“Korea” thereafter). This cross-country comparison is informed by key differences in contextual characteristics relating to gender in these countries such as cultural norms surrounding gender roles and workplace policies; however, we acknowledge that variation in gender-related experiences within a country (e.g., by race/ethnicity and socioeconomic status) may also shape the association between work expectations and mental health. We hypothesized that higher work expectations will be associated with lower likelihood of psychological distress and that this association will be stronger among men than among women. Further, differences in the association between work expectations and psychological distress across men and women will differ between the US and Korea.

## METHODS

### Data source and study population

Data came from two population-based longitudinal studies of aging conducted in the US and Korea, including the Health and Retirement Study (HRS, n∼20,000 respondents ages 51+ surveyed biennially since 1992) and the Korean Longitudinal Study of Aging (KLoSA, n∼10,000 respondents ages 45+ surveyed biennially since 2006). Both the HRS and KLoSA employ a complex, multi-stage study design that enrolls refreshment cohorts (every six years in the HRS and at the 2014 wave in KLoSA) to remain representative of the contemporaneous older adult population. These harmonized studies provide a unique opportunity to investigate the association between the expectations for late-life employment and psychological distress among older men and women in the US and Korea. Additional details on the HRS and KLoSA are described elsewhere (Joon et al., 2007; Sonnega et al., 2014).

This study focused on the Baby Boom cohorts born in the US (birth years: 1948 – 1965) and Korea (birth years: 1955 – 1963). The analytical sample included 16,367 non-proxy respondents (14,005 Americans and 2,362 Koreans) who had complete data on work expectations, psychological distress, and all potential covariates for at least one interview wave from 2006 to 2018. These interview waves were chosen because 2006 was the first interview wave of KLoSA, and 2018 was the latest available interview wave prior to the COVID-19 pandemic. While the pandemic may affect the association between work expectations and psychological distress, its impact is outside the scope of the present study.

Supplementary Figure 1 details the analytic sample selection process for the two cohorts. Differences between the analytic sample and the entire sample of Baby Boomers who were interviewed between 2006 and 2018 but excluded due to missing data on key variables of interest are summarized in Supplementary Table 1. Most relevant to this investigation, the excluded respondents were less likely to be working compared to the analytic sample.

The HRS and KLoSA are approved by the research ethics review committees in their respective countries, and all respondents are provided written informed consent. This analysis used only publicly available data (https://g2aging.org) and was exempt from human subject regulation.

### Work expectations

Questions on work expectations were asked at every interview wave in both the HRS and KLoSA; however, there are several key differences between the two datasets. First, the KLoSA assessed work expectations by a single question asking the respondents about their subjective likelihood of working “at the present job in the next 5 years.” Specifically, respondents younger than 50 were asked about the likelihood of working after age 55; respondents aged 50-54 were asked about working after age 60, respondents aged 55-59 were asked about working after age 65; and finally, respondents aged 60+ were asked about the likelihood of working in the next 5 years. On the other hand, the HRS assessed work expectations by three questions asking the respondents about the subjective likelihood of “working full-time” after age 62, 65, and 70.

These questions were then used to create a new variable based on the respondents’ age: expectations of working after age 62 were used for respondents aged 56 or younger; expectations of working after age 65 for those aged 57-62; and expectations of working after age 70 for those older than age 62. Overall, this variable assessed the likelihood of working full-time in the next 5 years, on average. Second, responses were reported on a probability scale of 0 to 100 at the interval of 10 in KLoSA, whereas responses were on a continuous scale from 0 to 100 in the HRS. Finally, work expectations were assessed only among respondents who were working at the time of assessment in the KLoSA; in contrast, the HRS assessed work expectations in all respondents irrespective of their working status. This difference in the assessment of work expectations informed the basis of the sensitivity analyses described below.

Work expectations were first modeled as a continuous time-varying exposure, and responses were rescaled to 0 to 10 for a more meaningful interpretation of regression coefficients (i.e., a one-unit increase in the scaled work expectation is equivalent to a ten-unit increase in the original variable). Additionally, reflecting the multi-modal distribution of this variable (e.g., clusters at tens and at 0%, 50% and 100%), a categorical version of work expectation (operationalized as 0-10%, 11-40%, 41-60%, 61-89%, 90-100%) to explore potential non-linear association between work expectations and psychological distress, with the category of 41-60% used as the reference to reflect “50/50 chance” uncertainty.

### Psychological distress

Psychological distress was assessed by the Center for Epidemiologic Studies Depression Scale (CES-D) on all respondents at every interview wave. The CES-D is originally a 20-item screening instrument that assesses four domains associated with distress symptoms (i.e., depressed effect, positive effect, somatic symptoms, and interpersonal relations) during the past week (Radloff, 1977). Although the HRS and KLoSA both derive their psychological distress measures from the CES-D, the HRS uses an 8-item scale, each recorded as yes/no (CES-D 8; scores range from 0 to 8). In contrast, the KLoSA uses a 10-item scale, each recorded on a 4-point Likert scale ranging from “very rarely (less than 1 day)” to “almost always (5 to 7 days)” (CES-D 10; scores range from 0 to 30). Specific items in CES-D 8 and CES-D 10 are summarized in Supplementary Table 2. Previous studies have shown that CES-D 8 and CES-D 10 have comparable psychometric properties (O’Halloran et al., 2014), acceptable internal consistency and test-retest reliability in older adults (Cronbach alpha is 0.78 – 0.84 for CES-D 8 and 0.72 – 0.80 for CES-D 10) (Cheng & Chan, 2005; Dang et al., 2020).

The CES-D summary scores were constructed by summing all symptoms, reverse-coding items on positive effects (e.g., “happiness” and “enjoying life”); higher scores indicated more severe symptomatology. To facilitate the comparison between the HRS and KLoSA, summary scores were then dichotomized to classify cases of psychological distress using threshold scores of ≥4 symptoms for CES-D 8 (HRS) and ≥10 for CES-D 10 (KLoSA). These thresholds have been shown to be equivalent to a threshold of 16 or more symptoms on the original 20-item CES-D (Steffick, 2000) and demonstrated acceptable sensitivity and specificity for distress symptoms in previous literature (Cheng & Chan, 2005; Dang et al., 2020).

### Covariates

All models adjusted for covariates related to demographic and health characteristics that have been associated with work expectations and psychological distress in prior work.

Demographics included age at baseline (centered at each sample’s mean), years of follow-up, education (less than lower secondary education, upper secondary & vocational training, tertiary education), marital status (married, not married), minority status, household income, total wealth, and employment status. Minority status (yes/no) was indicated as racial/ethnic minority group for the US (e.g., non-Hispanic Black, Hispanics, and others) and rural residential status for Korea. Household income and total wealth were categorized into quartiles based on baseline data. Employment status was categorized into full-time, part-time, self-employed, unemployed, partly retired, retired, disabled, and not in the labor force. Employment status was not included in models for Korean respondents as a covariate because work expectations were collected only among working respondents in KLoSA. Health characteristics included self-rate health (poor/fair, good, very good/excellent) and medical morbidity, indicated by having at least one chronic condition among hypertension, diabetes, cancer, chronic lung disease, heart problem, stroke, and arthritis. All covariates were time-varying as a function of interview wave except for race/ethnicity and baseline age.

### Statistical approach

Initially, baseline demographic and health characteristics for men and women were compared using Wilcoxon rank sum tests (comparing means) and Kruskal-Wallis tests (comparing medians) for continuous variables and chi-squared tests for categorical variables.

Mixed effects logistic regressions with random intercept and random slope of year were used to examine the association between work expectations and psychological distress over the 12-year period. This modeling approach accounts for the clustered structure in the data (i.e., within-person over time) and the time-varying nature of work expectations and psychological distress. It also incorporates information from all assessments rather than only those observations with complete follow-up over the study period (Hardin & Hilbe, 2018). The inclusion of random intercept term accounted for the heterogeneity in the baseline psychological distress; likelihood ratio test of the random intercept model vs. the model without was highly significant (p<0.0001). Temporal trends in psychological distress were modeled by the inclusion of both a fixed effect and a random slope for years of follow-up. The fixed effect term of year allows each respondent to have a different length of follow-up time, and the random slope term of year indicates a different rate of change in distress status over time. Models with interaction terms between years of follow-up and baseline age as well as between years of follow-up and work expectations were explored; however, these interaction terms were not statistically significant and thus removed from the analysis. All models were fit separately for men and women and by country (US, Korea).

Differences in the association between work expectations and psychological distress across men and women were tested by including an interaction term between self-reported gender and work expectations in the full sample of both men and women. Plots of average marginal predicted probabilities for psychological distress were displayed to visualize the findings. Lastly, since there is no consensus on CES-D threshold scores, additional analyses were conducted using alternative thresholds (≥3, ≥5 symptoms for CES-D 8 and ≥11, ≥12 symptoms for CES-D 10) to classify cases of psychological distress (Cheng & Chan, 2005; Dang et al., 2020). Statistical analyses were performed in the R environment, version 4.2.2 (2022).

Significance tests were evaluated two-sided at *p*<0.05. Mixed effects logistic regressions were fit via *glmer*, which estimates maximum likelihood by Laplace approximation. Marginal mean predicted probabilities were estimated with R package *ggeffects*, version 1.5.0.

## RESULTS

### Descriptive statistics

Table 1 shows the baseline characteristics of men and women in the US and Korea. In both countries, women had higher CES-D scores and lower mean expectations of working in the next 5 years than men. They were also less likely to be married, had lower educational attainment, had lower household income and wealth, and were less likely to be working (all *p*<0.05).

**Table 1.**
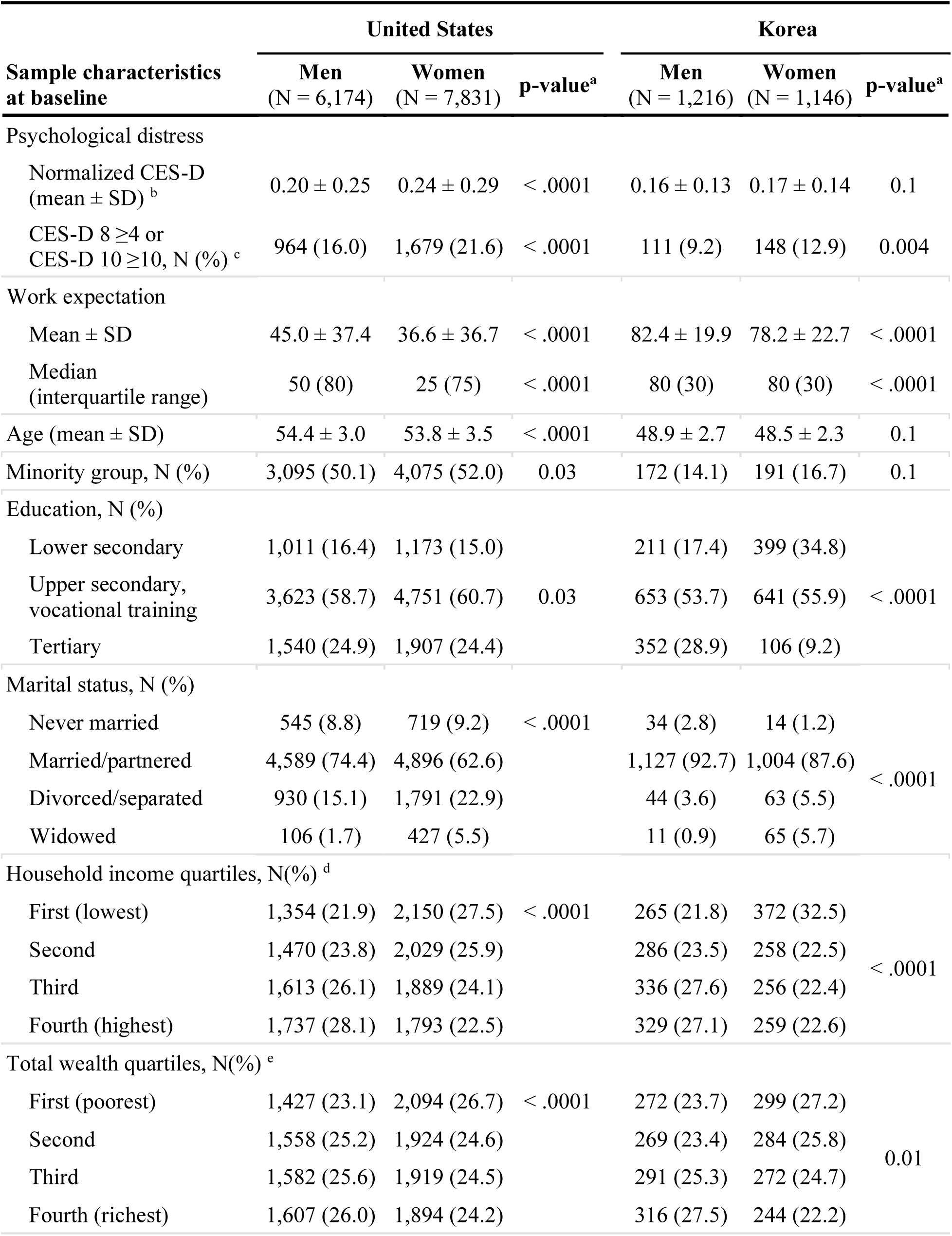
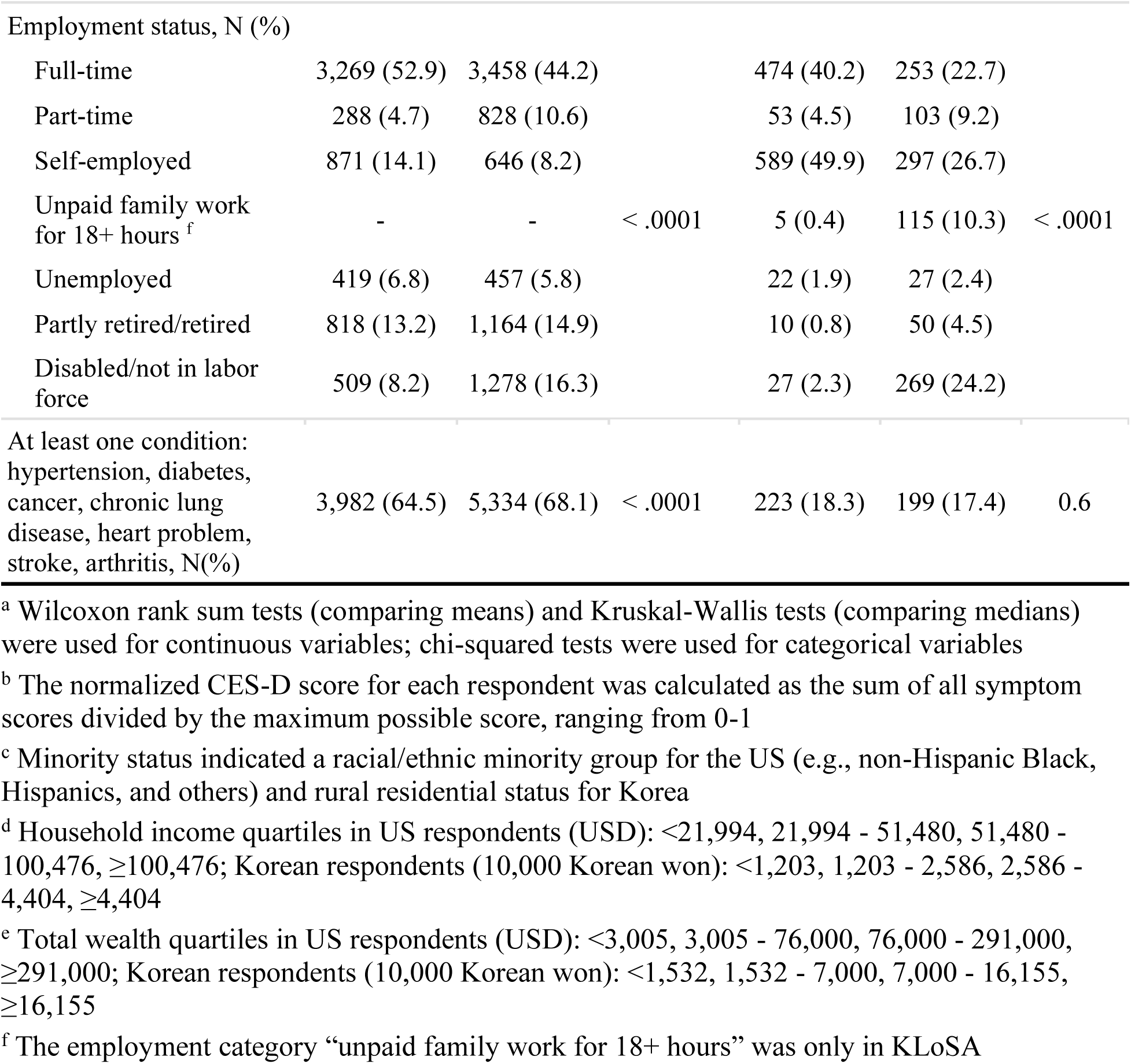
Sample characteristics of the United States and Korean respondents at baseline.

### Continuous work expectations and psychological distress among men and women

As shown in Table 2, in both countries, higher expectations of working in the next 5 years were associated with lower odds of psychological distress after adjusting for demographic and health characteristics. However, this inverse association was of a weaker magnitude in the US; moreover, the inverse association between expectations and distress were present among women (odds ratio (OR)_women_= 0.96, 95% confidence interval (CI): 0.95, 0.98 for every 10% increase in work expectation) but not men (OR_men_= 0.99 [95%CI=0.96, 1.01]) in the US (although this gender difference was not statistically significant (p=0.21)). In Korea, the inverse association was significantly stronger among men (OR_men_= 0.82 [95%CI=0.78, 0.87) compared with women (OR_women_= 0.88 [95%CI=0.83, 0.93]; p=0.02). Figure 1 illustrates these findings, showing that, in the US, higher work expectations were associated with slightly smaller predicted probabilities among women, but no association were observed among men (Figure 1A). In Korea, work expectations were also inversely associated with psychological distress, a relationship that was more pronounced among men (Figure 1B).

**Figure 1.**
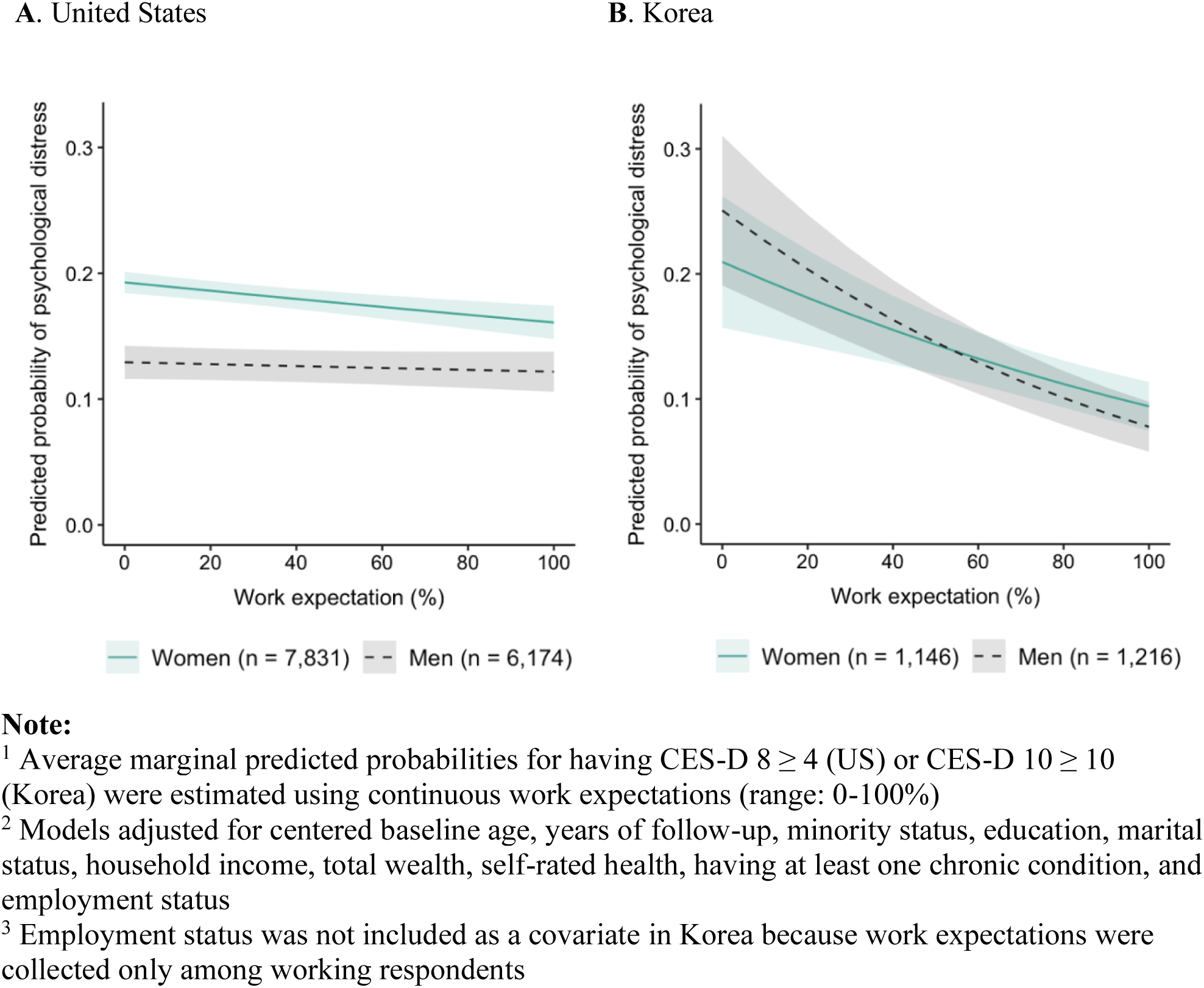
Average marginal predicted probability of psychological distress for Baby Boom men and women in the United States and Korea from 2006 to 2018

**Table 2.** Odd ratios and 95% confidence intervals for the association between work expectations and psychological distress by gender in the United States and Korea.

|  | Odds ratio (95% confidence interval) of psychological stress <sup>a</sup> |  |  |  |
| --- | --- | --- | --- | --- |
|  | United States |  | Korea <sup>b</sup> |  |
|  | Men<br>(N = 6,174) | Women<br>(N = 7,831) | Men<br>(N = 1,216) | Women<br>(N = 1,146) |
| <u>Continuous, scaled to 0-10%</u> |  |  |  |  |
| Effect estimate | 0.99<br>(0.96, 1.01) | 0.96<br>(0.95, 0.98)* | 0.82<br>(0.78, 0.87)* | 0.88<br>(0.83, 0.93)* |
| *p-value comparing ORs*<br>*in men and women* | 0.21 |  | **0.02** |  |
| <u>Categorical, reference = 41 - 60%</u> |  |  |  |  |
| 0 - 10% | 1.12<br>(0.88, 1.43) | 1.29<br>(1.10, 1.52)* | 1.58<br>(0.73, 3.42) | 1.10<br>(0.47, 2.57) |
| 11 - 40% | 0.89<br>(0.68, 1.17) | 0.99<br>(0.81, 1.20) | 1.52<br>(1.00, 2.34)* | 1.76<br>(1.12, 2.76)* |
| 41 - 60% | Ref | Ref | Ref | Ref |
| 61 - 89% | 0.78<br>(0.59, 1.04) | 0.96<br>(0.78, 1.18) | 0.64<br>(0.48, 0.86)* | 0.66<br>(0.48, 0.91)* |
| 90 - 100% | 1.12<br>(0.84, 1.49) | 0.96<br>(0.78, 1.18) | 0.42<br>(0.30, 0.60)* | 0.66<br>(0.46, 0.94)* |

<sup>a</sup> Odds ratios were calculated for having CES-D 8 ≥ 4 (United States) or CES-D 10 ≥ 10 (Korea). Models adjusted for centered baseline age, years of follow-up, minority status, education, marital status, household income, total wealth, self-rated health, having at least one chronic condition, and employment status
<sup>b</sup> Employment status was not included as a covariate in Korea because work expectations were collected only among working respondents
\* $p < 0.05$

### Categorical work expectations and psychological distress in men and women

Next, to explore potential non-linear relationships, the exposure of work expectations were modeled as a categorical variable, with the reference group of “uncertain” (expectations of 41-60%). Table 2 shows that, in the US, using this categorization there was no significant association between work expectations and psychological distress among men; however, women who reported low expectations of working (i.e., 0-10%) had significantly higher odds of distress relative to those who were uncertain (OR_women_= 1.29 [95%CI=1.10, 1.52]). In Korea, respondents with lower expectations of working also had higher odds for psychological distress, though this inverse association was only statistically significant when comparing respondents who reported expectations of 11-40% with those with uncertain expectations (OR_men_= 1.52 [95%CI=1.00, 2.34] and OR_women_= 1.76 [95%CI=1.12, 2.76]). Additionally, higher expectations of working (between 61-100%) were associated with lower odds of distress in both Korean men (OR_61-89%_= 0.64 [95%CI=0.48, 0.86] and OR_90-100%_= 0.42 [95%CI=0.30, 0.60]) and women (OR_61-89%_= 0.66 [95%CI=0.48, 0.91] and OR_90-100%_= 0.66 [95%CI=0.46, 0.94]).

Finally, as shown in Supplementary Tables 3-4, using alternative threshold scores of CES-D for classifying psychological distress similarly found a stronger inverse association between work expectations and psychological distress among Korean men compared to women.

In the US, results were consistent that the inverse association between work expectations and psychological distress was found among women but not men.

## DISCUSSION

The goal of this longitudinal study was to explore the relationships between gender, work, and aging among the Baby Boom generation using an international comparative approach. The primary findings of this study are three-fold. First, we found that higher work expectations were robustly associated with lower odds of psychological distress among both men and women of the Baby Boom generation. Second, this inverse association was more modest among US respondents compared to their Korean counterparts. Finally, the inverse association between work expectations and psychological distress was stronger among Korean men compared to women; in contrast, there were no gender differences among US respondents. Collectively, these findings suggest that there are multifaceted processes shaping gender differences in the association between work expectations and psychological distress among Baby Boomers. This study illustrates the ways in which contextual factors, such as age cohort, gender, and social norms, contribute to the links between work and mental health in later life.

In Korea, the robust inverse association between work expectations and psychological distress may reflect the employment practices and pension policies that are salient to the mental health of older workers. One example of this is the finding that older Korean adults, overall, expect to work longer than their US counterparts (Table 1). The expectations of working longer expressed by older Korean workers could be driven by financial necessity as a consequence of a weak social safety net. In Korea, the Basic Pension system provides pension benefits for older adults with earnings in the bottom 70% of the income distribution. However, the gross pension replacement rates are relatively low; in 2022, this rate was estimated at 31% for a full-career worker [from age 22 to 59] with an average wage compared to an international average of 50% (OECD, 2019). Moreover, the current pension system largely excludes non-regular employment such as self-employed, part-time, or temporary work. These types of non-regular forms of work do not typically offer pension benefits or pay contributions to public pensions (Kuhlmann & Nullmeier, 2022). However, these work arrangements are especially prevalent among older Koreans, accounting for nearly half of salaried workers over age 60 (OECD, 2019). Given the widespread practice of “honorary” retirement (in which Korean firms incentivize older workers to retire, often in the mid-50s age range), many older Koreans involuntarily retire early from their career jobs before they are qualified to receive pension benefits [at age 60] (Cho & Kim, 2005). These older adults often seek new employment in non-regular jobs to earn or supplement their living expenses (OECD, 2019). In the context of such precarious work environment, the association between work expectations and distress could reflect the distress around securing stable employment among older Koreans.

In the US, work expectations were only modestly inversely associated with psychological distress among women. While, overall, the inverse association between expectations and distress is consistent with findings from our prior study examining work expectations and indicators of depression in the HRS (Mezuk et al., 2022), the magnitude is smaller. This difference in magnitude may stem from differences in measurement: in the present study, psychological distress was measured using the CES-D, which only assesses current (i.e., past week) symptoms. In contrast, our prior study used the Composite International Diagnostic Interview (CIDI), a fully structured interview based on the Diagnostic and Statistical Manual of Mental Disorders-IV criteria for major depressive episodes, to assess depressive symptoms over the past 12 months.

Given that planning for future employment or an eventual exit from the workforce typically takes place over an extended period of months or years (Topa et al., 2018), the CIDI may be better suited to assessing psychological distress associated with long-term retirement planning.

Furthermore, it is possible that the effect of gender on the expectation-distress association is more complex and may be influenced by factors such as family/spousal relationships (Fisher et al., 2016). Such a hypothesis is supported by findings from previous studies on this topic; for example, a recent analysis found that marriage/partnership and gender roles moderated the association between employment stability and mental health (Cortès-Franch et al., 2018).

### Exploring cohort differences using a cross-national approach

The key finding from this study is that while the association between work expectations and psychological distress was stronger among Korean men compared to women, there was no significant gender difference among their US counterparts. This cross-country finding should be interpreted from the life course perspective, which emphasizes how temporal (e.g., experiences earlier in life) and contextual (e.g., societal norms about gender roles) factors shape work and mental health in later life. The Baby Boom cohort grew up during a period when both countries underwent significant economic changes. Specifically, the US Baby Boomers entered the workforce during post-World War II period when the US experienced rapid economic growth (Pruitt, 2020). On the other hand, the Korean Baby Boomers spent their childhood in the aftermath of the Korean War and entered adulthood when Korea underwent a phenomenal transformation from a war-ravaged, impoverished country to an industrialized, highly developed economy (Park, 2019). These periods of economic growth resulted in an abundance of employment opportunities and increasing labor force participation, especially among workers from the Baby Boom cohort (Poulos & Nightingale, 1997). While both genders have strong attachments to the workforce (Fry, 2019), labor force participation, particularly in women, is strongly dependent on the social norms and expectations regarding gender roles within each culture. Compared to the US, Korean culture is influenced by Confucian values which maintain a rigid gender norm and clear division of labor for men and women (Fauser & Kim, 2023). Thus, Korean women are more likely to exit the labor force for caregiving responsibilities or engage in non-regular forms of work after childbirth than men (Kim, 2022). Indeed, data from the Korean Labor and Income Panel Study showed that the majority of Korean women held more precarious, non-regular work positions such as unpaid family work, self-employment, and low pay, temporary jobs; only one-fifth were employed in regular jobs after childbirth (Fauser & Kim, 2023). These cross-culture differences in societal norms for men’s and women’s roles influence the gender differences in selection and/or retention in the workforce, which in turn contributes to the gendered patterns in the association between work expectations and psychological distress in the US and Korea.

Overall, findings illustrate a complex processes that contribute to the retirement process among men and women. Epidemiologic research in the area of work and health has long recognized the distinction between sex (i.e., as a biological construct based on genetics, anatomical and physiological differences) and gender (i.e., social construct based on societal and cultural norms for men and women) (Smith & Koehoorn, 2016). Such distinction seeks to differentiate the biological aspect from the social and cultural aspects of gender. However, sex and gender often interact to influence behaviors, including retirement decisions (Springer et al., 2012). This study provides further evidence to support such interaction, showing that the differences in the association between work expectations and psychological distress varied not only by biological sex (male/female) but also by country contexts which guide the societal expectations for men’s and women’s roles. It emphasizes the importance of integrating both biological and social risk factors to inform mental health efforts supporting older men and women during the retirement transition.

Findings should be interpreted considering study limitations and strengths. First, the analytical sample was more likely to be working than the Baby Boom cohort in general. This was due, in part, to the fact that the KLoSA only asked about work expectations among respondents who were working at the time of assessment. Therefore, findings may not extend to older adults who were not currently working (e.g., were unemployed or temporarily laid off). These employment transitions are more prevalent among women, who may work intermittently or take time off work for caregiving responsibilities (Richardson, 1999). Second, possible reverse causation may confound the study results given that psychological distress could affect the expectations and ability to work. Future studies could address the potential of reverse causality by including data on early life experiences and prior diagnoses of psychological distress. Third, the study did not have data on gender attitudes and individual perceptions of work and family responsibilities. Such data is particularly important in cross-culture contexts where work may have different meanings in Korean and American cultures (e.g., a financial means to support family vs. career achievement). Lastly, psychological distress was assessed by the CES-D rather than clinical interviews and as such this may not reflect clinically significant diagnoses. Future studies could use more robust depression measures as well as examine other mental health outcomes (e.g., anxiety) and related outcomes (e.g., loneliness).

Despite limitations, the study has several strengths. To the best of our knowledge, this study is among the first to examine the gender difference in the association between work expectations and psychological distress among older adults. While most studies on work and mental health have been conducted in Western countries, this study situated the association between work expectations and distress within the US and Korea. This cross-country comparison provides important insights into how societal norms and expectations about gender roles within a given country influence individuals’ decisions about work and its impacts on psychological distress. Future research can expand this analysis to other countries to gain a more comprehensive, international perspective of how work expectations relate to psychological distress among men and women. Insights from such comparisons help to inform culturally appropriate interventions to support the mental health needs of an increasingly diverse population of older workers. Finally, the study sample came from large population-based studies with rich data on employment and distress measures, which enhances the study’s validity.

## CONCLUSION

The study found an inverse association between work expectations and psychological distress among both men and women of the Baby Boom generation. This inverse association was stronger among Korean men compared to women; in contrast, there were no gender differences among American respondents. Findings highlight the multifaceted processes, including age cohort, gender, and social norms, that shape work and mental health in later life.

## Data Availability

All data produced in the present work are described in the manuscript.

## ACKNOWLEDGMENTS

We have no conflict of interest to declare. The HRS and KLoSA are publicly available to researchers at https://hrs.isr.umich.edu and https://survey.keis.or.kr/eng/klosa/klosa01.jsp.

---

**Supplementary Figure 1.**
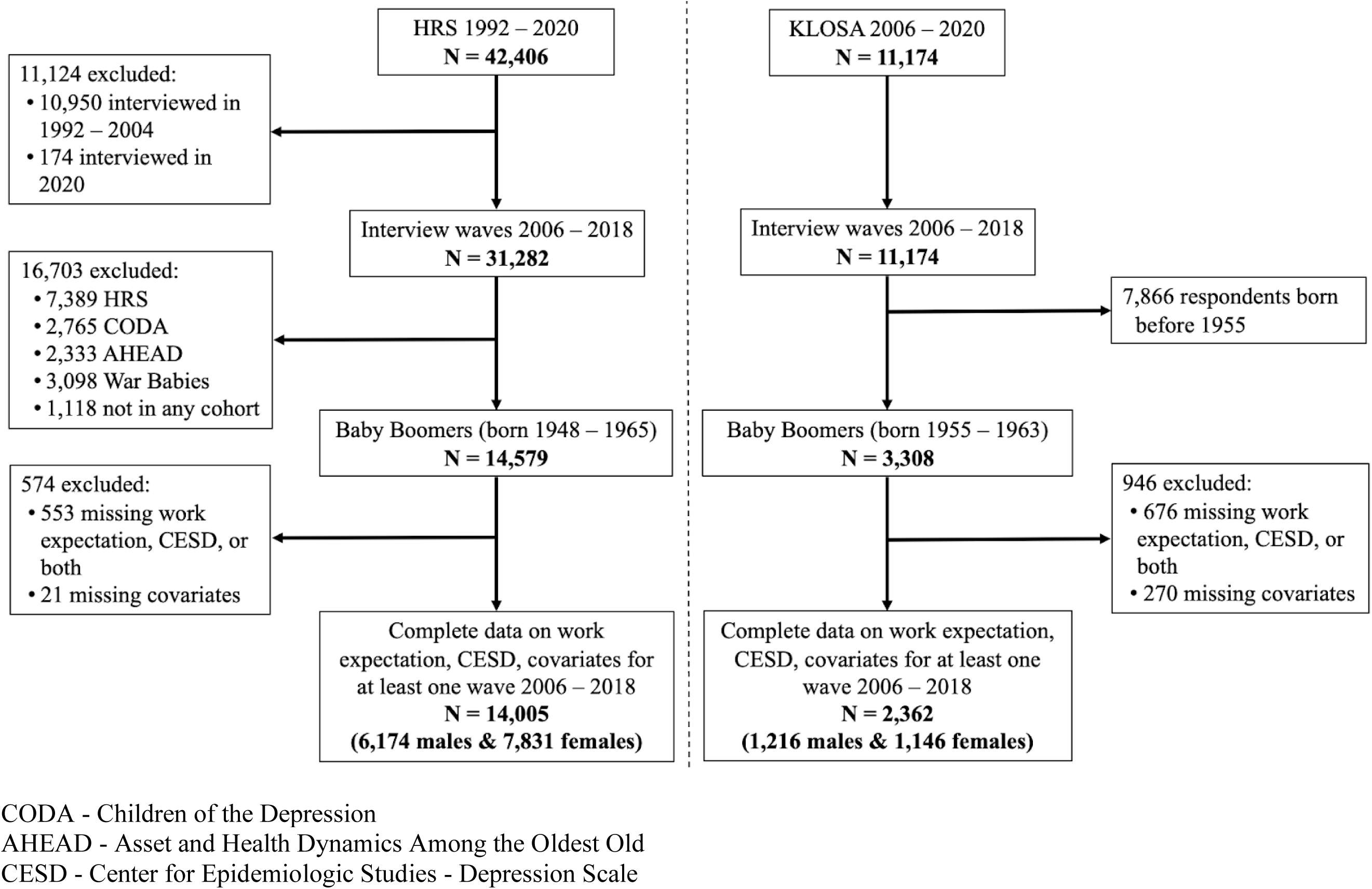
Selection criteria of the analytical samples in the Health and Retirement Study (HRS) and Korean Longitudinal Study of Aging (KLoSA)

**Supplementary Table 1.** Characteristics of included and excluded Baby Boomers who were interviewed between 2006 and 2018 at the first assessment.

| Characteristics at baseline | United States |  |  | Korea |  |  |
| --- | --- | --- | --- | --- | --- | --- |
|  | Included<br>(N =14,005) | Excluded<br>(N = 574) | p-value<sup>a</sup> | Included<br>(N = 2,362) | Excluded<br>(N = 946) | p-value<sup>a</sup> |
| Psychological distress |  |  |  |  |  |  |
| Normalized CES-D<br>(mean ± SD) <sup>b</sup> | 0.22 ± 0.27 | 0.25 ± 0.28 | 0.07 | 0.17 ± 0.13 | 0.19 ± 0.17 | 0.004 |
| CES-D 8 ≥4 or<br>CES-D 10 ≥10, N (%) | 2,643 (19.2) | 70 (19.7) | 0.8 | 259 (11.0) | 151 (16.1) | < .0001 |
| Work expectation |  |  |  |  |  |  |
| Mean ± SD | 40.5 ± 37.3 | 32.2 ± 36.3 | 0.2 | 80.7 ± 21.2 | 70.4 ± 21.5 | < .0001 |
| Median<br>(interquartile range) | 40 (80) | 20 (50) | 0.2 | 80 (30) | 70 (30) | < .0001 |
| Age (mean ± SD) | 54.0 ± 3.3 | 52.5 ± 4.8 | < .0001 | 48.7 ± 2.5 | 49.9 ± 2.7 | < .0001 |
| Female, N (%) | 7,831 (55.9) | 281 (49.0) | 0.001 | 1,146 (48.5) | 701 (74.1) | < .0001 |
| Minority group, N (%) <sup>c</sup> | 7,170 (51.2) | 281 (49.5) | 0.4 | 363 (15.4) | 79 (8.4) | < .0001 |
| Education, N (%) |  |  |  |  |  |  |
| Lower secondary | 2,184 (15.6) | 122 (21.3) |  | 610 (25.8) | 201 (21.3) |  |
| Upper secondary,<br>vocational training | 8,374 (59.8) | 324 (56.6) | 0.001 | 1,294 (54.8) | 536 (56.8) | 0.02 |
| Tertiary | 3,447 (24.6) | 126 (22.0) |  | 458 (19.4) | 207 (21.9) |  |
| Marital status, N (%) |  |  |  |  |  |  |
| Never married | 1,256 (9.0) | 22 (3.9) |  | 48 (2.0) | 29 (3.1) |  |
| Married/partnered | 9,485 (67.8) | 490 (85.8) | < .0001 | 2,123 (90.2) | 853 (90.2) | 0.09 |
| Divorced/separated | 2,721 (19.4) | 49 (8.6) |  | 107 (4.5) | 45 (4.8) |  |
| Widowed | 533 (3.8) | 10 (1.8) |  | 76 (3.2) | 19 (2.0) |  |
| Household income quartiles, N(%) <sup>d</sup> |  |  |  |  |  |  |
| First (lowest) | 3,504 (25.0) | 146 (25.4) |  | 637 (27.0) | 334 (35.4) |  |
| Second | 3,499 (25.0) | 132 (23.0) | 0.2 | 544 (23.0) | 151 (16.0) | < .0001 |
| Third | 3,502 (25.0) | 132 (23.0) |  | 592 (25.1) | 192 (20.4) |  |
| Fourth (highest) | 3,500 (25.0) | 164 (28.6) |  | 588 (24.9) | 266 (28.2) |  |
| Total wealth quartiles, N(%)<sup>e</sup> |  |  |  |  |  |  |
| First (poorest) | 3,521 (25.1) | 128 (22.3) |  | 571 (25.4) | 256 (30.2) |  |
| Second | 3,482 (24.9) | 140 (24.4) | 0.3 | 553 (24.6) | 170 (20.0) | < .0001 |
| Third | 3,501 (25.0) | 145 (25.3) |  | 563 (25.1) | 173 (20.4) |  |
| Fourth (richest) | 3,501 (25.0) | 161 (28.0) |  | 560 (24.9) | 250 (29.4) |  |
| Employment status, N (%) |  |  |  |  |  |  |
| Full-time | 6,727 (48.0) | 232 (40.4) |  | 727 (31.7) | 3 (0.5) |  |
| Part-time | 1,116 (8.0) | 29 (5.1) |  | 156 (6.8) | 2 (0.3) |  |
| Self-employed | 1,517 (10.8) | 76 (13.2) | < .0001 | 886 (38.6) | 11 (1.7) | < .0001 |
| Unpaid family work<br>for 18+ hours<sup>f</sup> | - | - |  | 120 (5.2) | 0 (0) |  |
| Unemployed | 876 (6.3) | 20 (3.5) |  | 49 (2.1) | 25 (3.9) |  |
| Partly retired/retired | 1,982 (14.2) | 55 (9.6) |  | 60 (2.6) | 124 (19.3) |  |
| Disabled/not in labor<br>force | 1,787 (12.7) | 162 (28.2) |  | 296 (12.9) | 477 (74.3) |  |
| At least one condition:<br>hypertension,<br>diabetes, cancer,<br>chronic lung disease,<br>heart problem, stroke,<br>arthritis, N(%) | 9,316 (66.5) | 344 (59.9) | 0.001 | 422 (17.9) | 175 (18.5) | 0.7 |

<sup>a</sup> Wilcoxon rank sum tests (comparing means) and Kruskal-Wallis tests (comparing medians) were used for continuous variables; chi-squared tests were used for categorical variables
<sup>b</sup> The normalized CES-D score for each respondent was calculated as the sum of all symptom scores divided by the maximum possible score, ranging from 0-1
<sup>c</sup> Minority status indicated a racial/ethnic minority group for the US (e.g., non-Hispanic Black, Hispanics, and others) and rural residential status for Korea
<sup>d</sup> Household income quartiles in US respondents (USD): <21,994, 21,994 - 51,480, 51,480 - 100,476, ≥100,476; Korean respondents (10,000 Korean won): <1,203, 1,203 - 2,586, 2,586 - 4,404, ≥4,404
<sup>e</sup> Total wealth quartiles in US respondents (USD): <3,005, 3,005 - 76,000, 76,000 - 291,000, ≥291,000; Korean respondents (10,000 Korean won): <1,532, 1,532 - 7,000, 7,000 - 16,155, ≥16,155
<sup>f</sup> The employment category “unpaid family work for 18+ hours” was only in KLoSA

**Supplementary Table 2.** Items included in the 8-item (Health and Retirement Study, HRS) and 10-item (Korean Longitudinal Study of Aging, KLoSA) Center for Epidemiologic Studies – Depression Scale.

| CES-D 8 |  | CES-D 10 |
| --- | --- | --- |
| **In the past week, did you...** |  |  |
| **Item 1** | ... feel depressed? | ... feel depressed? |
| **Item 2** | ... feel everything was an effort? | ... feel everything was an effort? |
| **Item 3** | ... sleep was restless? | ... sleep was restless? |
| **Item 4** | ... feel happy? | ... feel happy? |
| **Item 5** | ... feel lonely? | ... feel lonely? |
| **Item 6** | ... enjoy life? | ... enjoy life? |
| **Item 7** | ... could not get going? | ... could not get going? |
| **Item 8a<sup>a</sup>** | ... feel sad? | ... feel sad? |
| **Items available only in CES-D 10 (KLoSA)** |  |  |
| *Wave 1 - 4 of KLoSA (2006 - 2012)<sup>b</sup>* |  |  |
| **Item 8b** | - | ... has trouble concentrating? |
| **Item 9** | - | ... feel bothered by things? |
| **Item 10** | - | ... feel afraid? |
| *Wave 5 - 7 of KLoSA (2014 - 2018)<sup>c</sup>* |  |  |
| **Item 9** | - | ... feel that people were unfriendly? |
| **Item 10** | - | ... feel that people disliked you? |

<sup>a</sup> Wave 1-4 of KLoSA did not include question asking whether the respondent felt sad (item 8a)
<sup>b</sup> Wave 1-4 of KLoSA included three additional questions asking whether the respondents had trouble concentrating, felt bothered by things, and felt afraid
<sup>c</sup> From wave 5 onward, KLoSA included all items asked in the HRS and two additional questions asking whether the respondents felt that people were unfriendly and people disliked them
<sup>d</sup> CES-D 8 used yes/no responses with scores ranging from 0-8, whereas CES-D 10 was reported on a 4-point Likert scale (very rarely, sometimes, often, almost always) and ranged from 0-30

**Supplementary Table 3.** Odd ratios and 95% confidence intervals for the association between work expectations and psychological distress by gender in the United States using alternative threshold scores for CES-D 8 (CES-D 8 ≥ 3, ≥ 5)

|  | Odds ratio (95% confidence interval) |  |
| --- | --- | --- |
|  | Men<br>(N = 6,174) | Women<br>(N = 7,831) |
| CES-D 8 ≥ 3 |  |  |
| <u>Continuous, scaled to 0-10%</u> |  |  |
| Effect estimate | 0.98 (0.96, 1.00)* | 0.98 (0.96, 0.99)* |
| *p-value comparing ORs in men and women* | 0.83 |  |
| <u>Categorical, reference = 41 - 60%</u> |  |  |
| 0 - 10% | 1.05 (0.87, 1.27) | 1.23 (1.07, 1.41)* |
| 11 - 40% | 0.89 (0.72, 1.11) | 1.17 (1.00 1.37)* |
| 41 - 60% | Ref | Ref |
| 61 - 89% | 0.76 (0.61, 0.94)* | 1.07 (0.90, 1.27) |
| 90 - 100% | 0.86 (0.69, 1.07) | 1.05 (0.89, 1.25) |
| CES-D 8 ≥ 5 |  |  |
| <u>Continuous, scaled to 0-10</u> |  |  |
| Effect estimate | 0.98 (0.95, 1.02) | 0.97 (0.95, 0.99)* |
| *p-value comparing ORs in men and women* | 0.40 |  |
| <u>Categorical, reference = 41 - 60%</u> |  |  |
| 0 - 10% | 1.19 (0.85, 1.68) | 1.24 (1.03, 1.49)* |
| 11 - 40% | 1.07 (0.73, 1.58) | 1.02 (0.82, 1.27) |
| 41 - 60% | Ref | Ref |
| 61 - 89% | 0.86 (0.57, 1.29) | 0.95 (0.75, 1.20) |
| 90 - 100% | 1.29 (0.86, 1.94) | 1.01 (0.80, 1.28) |

All models adjusted for centered baseline age, years of follow-up, minority status, education, marital status, household income, total wealth, self-rated health, having at least one chronic condition, and employment status
\* $p < .05$

**Supplementary Table 4.**
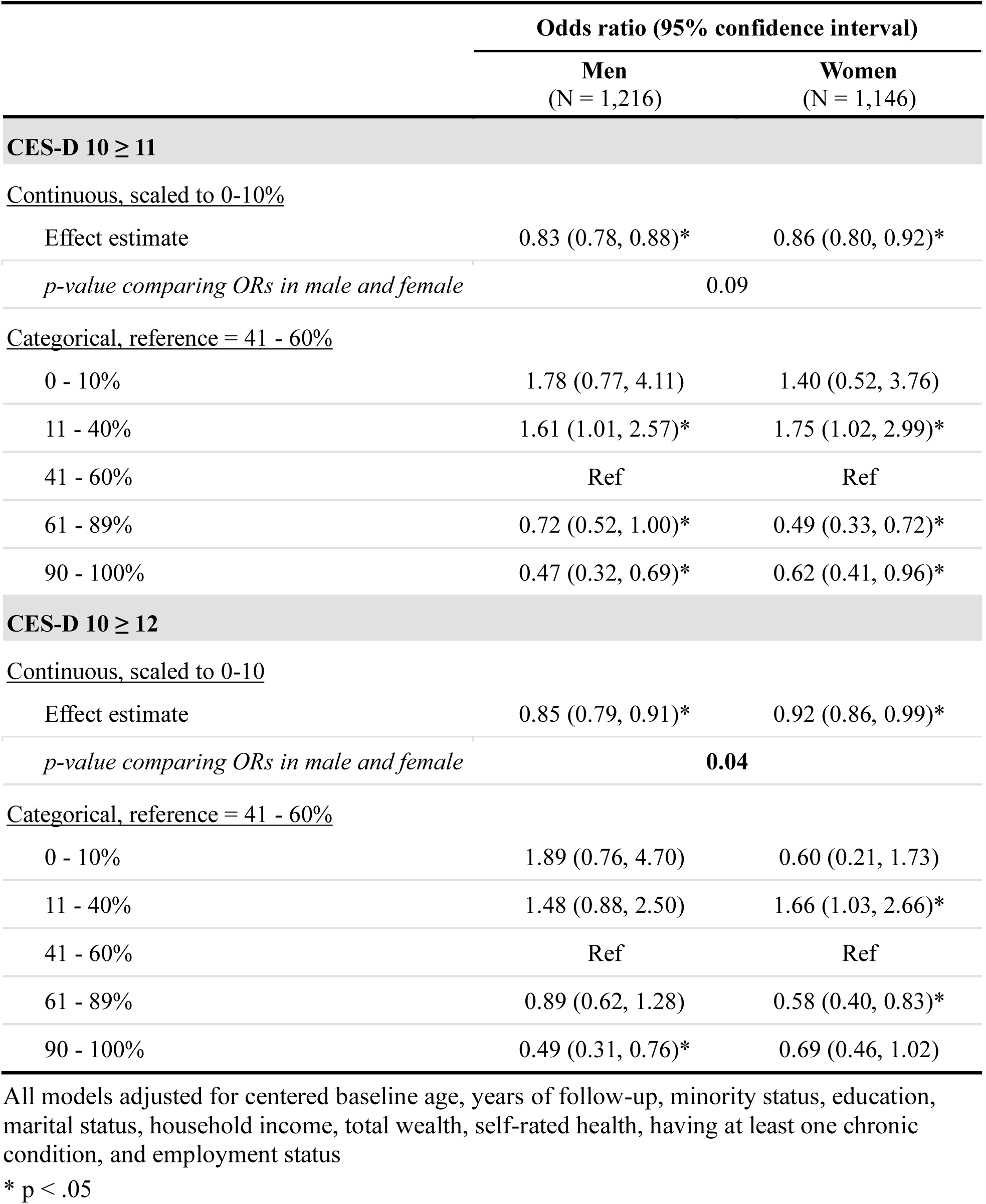
Odd ratios and 95% confidence intervals for the association between work expectations and psychological distress by gender in Korea using alternative threshold scores for CES-D 10 (CES-D 10 ≥ 11, ≥ 12)

|  | Odds ratio (95% confidence interval) |  |
| --- | --- | --- |
|  | Men<br>(N = 1,216) | Women<br>(N = 1,146) |
| CES-D 10 ≥ 11 |  |  |
| <u>Continuous, scaled to 0-10%</u> |  |  |
| Effect estimate | 0.83 (0.78, 0.88)* | 0.86 (0.80, 0.92)* |
| *p-value comparing ORs in male and female* | 0.09 |  |
| <u>Categorical, reference = 41 - 60%</u> |  |  |
| 0 - 10% | 1.78 (0.77, 4.11) | 1.40 (0.52, 3.76) |
| 11 - 40% | 1.61 (1.01, 2.57)* | 1.75 (1.02, 2.99)* |
| 41 - 60% | Ref | Ref |
| 61 - 89% | 0.72 (0.52, 1.00)* | 0.49 (0.33, 0.72)* |
| 90 - 100% | 0.47 (0.32, 0.69)* | 0.62 (0.41, 0.96)* |
| CES-D 10 ≥ 12 |  |  |
| <u>Continuous, scaled to 0-10</u> |  |  |
| Effect estimate | 0.85 (0.79, 0.91)* | 0.92 (0.86, 0.99)* |
| *p-value comparing ORs in male and female* | 0.04 |  |
| <u>Categorical, reference = 41 - 60%</u> |  |  |
| 0 - 10% | 1.89 (0.76, 4.70) | 0.60 (0.21, 1.73) |
| 11 - 40% | 1.48 (0.88, 2.50) | 1.66 (1.03, 2.66)* |
| 41 - 60% | Ref | Ref |
| 61 - 89% | 0.89 (0.62, 1.28) | 0.58 (0.40, 0.83)* |
| 90 - 100% | 0.49 (0.31, 0.76)* | 0.69 (0.46, 1.02) |

All models adjusted for centered baseline age, years of follow-up, minority status, education, marital status, household income, total wealth, self-rated health, having at least one chronic condition, and employment status
\* $p < .05$

## Notes

### Competing Interest Statement

The authors have declared no competing interest.

### Funding Statement

This study was supported by the National Institute of Mental Health (1R01MH128198-01) and National Institute on Aging (T32AG027708).

### Author Declarations

The study used only openly available human data that were originally located at https://g2aging.org.

